# Adaptive single-channel EEG artifact removal with applications to clinical monitoring

**DOI:** 10.1101/2021.10.19.21265197

**Authors:** Matteo Dora, David Holcman

## Abstract

**Objective:** Electroencephalography (EEG) has become very common in clinical practice due to its relatively low cost, ease of installation, non-invasiveness, and good temporal resolution. Portable EEG devices are increasingly popular in clinical monitoring applications such as sleep scoring or anesthesia monitoring. In these situations, for reasons of speed and simplicity only few electrodes are used and contamination of the EEG signal by artifacts is inevitable. Visual inspection and manual removal of artifacts is often not possible, especially in real-time applications. Our goal is to develop a flexible technique to remove EEG artifacts in these contexts with minimal supervision.

**Methods:** We propose here a new wavelet-based method which allows to remove artifacts from single-channel EEGs. The method is based on a data-driven renormalization of the wavelet components and is capable of adaptively attenuate artifacts of different nature. We benchmark our method against alternative artifact removal techniques.

**Results:** We assessed the performance of the proposed method on publicly available datasets comprising ocular, muscular, and movement artifacts. The proposed method shows superior performances on different kinds of artifacts and signal-to-noise levels. Finally, we present an application of our method to the monitoring of general anesthesia.

**Conclusions:** We show that our method can successfully attenuate various types of artifacts in single-channel EEG.

**Significance:** Thanks to its data-driven approach and low computational cost, the proposed method provides a valuable tool to remove artifacts in real-time EEG applications with few electrodes, such as monitoring in special care units.

## I. Introduction

**E**lectroencephalography (EEG) is a non-invasive procedure to monitor brain activity by recording electrical signal from electrodes placed on the scalp of a patient. Compact, portable and easy-to-use EEG devices are now routinely used for clinical needs. For example, EEG is employed to monitor the depth of general anesthesia [1], a method pioneered by the introduction of the bispectral index (BIS) in the 90s [2]. Today, more than ten EEG monitoring devices for anesthesia monitoring are available on the market [3]. Similar devices are used for automatic monitoring and scoring of sleep [4] or for evaluating neurological disorders in intensive care units (ICUs) [5], [6]. Outside of the clinical setting, ambulatory electroencephalography (AEEG) allows the acquisition of EEG data through a portable device which can be carried by the patient and can record up to 72 hours of activity. With the fast development of telemedicine, EEG recordings can even be sent in real-time for remote interpretation [7]. Tele-EEG systems are particularly useful for hospitals which do not dispose of resident neurophysiologists [7]–[9].

All these EEG monitoring applications are made possible by the little to no supervision needed to operate the device and the ease of installation even by non-technical staff. The downside of operating in these diverse and uncontrolled conditions is the possible contamination of the signal by extraneous sources. This is especially common for devices operating in intensive care units, epilepsy monitoring units, or the operating room [10]. Since signal intensity in scalp EEG is weak (typically 20 μV to 100 μV amplitude in adults [11]), even small variations of the electric signal can produce visible artifacts on the EEG traces. Examples are eye movements and blinks, muscular or cardiac activity, motion of electrodes or cables, skin perspiration, and interferences caused by electrical devices such as pacemakers [11], [12].

While artifacts can be recognized and sampled out by visual inspection when the analysis is performed offline by practitioners, they pose a major problem in monitoring devices which operate in real-time and unsupervised. In most cases artifacts can be detected quite reliably by localizing non-physiological high voltages or frequencies. Typically, clinical monitoring devices include some artifact detection and they obviate the problem by temporarily suspending the monitoring and resuming it only after the ratio between artifacted and clean signal drops below a given threshold. During anesthesia, this interruption deprives the medical staff of relevant information and may delay decision making. It is thus desirable to assure the continuity of monitoring even in the presence of artifacts.

Several methods have been proposed for EEG artifact removal. One of the most common techniques is blind source separation (BSS) via independent component analysis (ICA) [13], [14]. Based on the principle that brain activity is independent of artifactual sources, in ICA-based artifact removal the EEG signal is decomposed into a set of independent sources and the artifactual ones are selectively suppressed. This procedure is semi-automated as it requires identification of artifactual sources. The identification can be performed by visual inspection, but automated classification methods are also available [15], [16]. The quality of ICA decomposition is highly dependent on the preprocessing procedure [17]. Although good results can be achieved by applying high-pass (or band-pass) filters before ICA, this filtering procedure may also remove relevant information [17], [18]. Moreover, source separation techniques require multiple channels. This need constitutes a major limitation when working with EEG monitoring devices designed for operating rooms, which typically provide only one to four electrodes [3]. To overcome this issue, BSS algorithms are sometimes used in association with wavelet transform (WT) or empirical mode decomposition (EMD). The decomposition by WT or EMD allows to split a one-dimensional signal into multiple components which are then used as a multichannel input for BSS. Examples of this approach are WICA [19], EMD-ICA [20], and EMD-CCA [21]. However, one major drawback of source separation techniques such as ICA is that they cannot be applied on-line [14]. Although some real-time applications have been proposed by recurrently computing ICA on a sliding window [22], [23], no efficient on-line algorithm is today available. Moreover, the performances of ICA are affected by the length of the signal [24]: on one hand, a too short signal can prevent reliable separation of sources; on the other hand, when the signal is too long, the properties of the sources can change in time leading to improper isolation of the artifacts. These limitations make ICA and similar BSS-based methods unpractical in the context of real-time continuous monitoring.

An alternative to ICA are wavelet-based methods. The effectiveness of the wavelet transform (WT) at capturing time-frequency patterns at multiple scales has made it a common technique in various noise removal tasks, with applications ranging from images [25], audio [26], to physiological signals [27]–[29]. The WT decomposes a signal into multiple sets of coefficients representing time-frequency patterns of the signal. Denoising a signal is typically achieved by thresholding the wavelet coefficients, eliminating those corresponding to noise. Due to their flexibility, efficiency, and robustness, wavelet-based methods have been widely used for EEG artifact removal. Compared to blind source separation, the WT has a low computational cost, can be applied on-line, and does not require multiple channels. Wavelet-based methods can be very efficient at eliminating artifacts when the chosen wavelet basis allows good separation of the signal from noise [13]. Today, methods can combine wavelet decomposition with ICA [19], [30], [31].

In this article, we introduce a wavelet-based method called wavelet quantile normalization (WQN) which allows to eliminate transient artifacts from single-channel EEGs. The method consists in normalizing the wavelet coefficients of artifacted intervals, using temporally adjacent uncontaminated signal as a reference. This approach is relevant for continuous monitoring during general anesthesia and coma, where we expect a temporal continuity of the power distribution of the signal. The parameters used in the WQN method are estimated from the EEG signal itself with no prior human intervention. In this work, we consider the segmentation of artifactual epochs to be known. This requirement can be alleviated by using an appropriate artifact detection algorithm, similarly to those employed in BSS-based methods, making WQN artifact removal completely automated. Most EEG monitoring devices already include artifact detection algorithms so that automated analysis can be suspended when the signal quality is degraded.

This paper is organized as follows. In section II we introduce the notation and the discrete wavelet transform (II-A), we briefly present the classical wavelet denoising approaches which we used to benchmark our method (II-B), and finally we present the wavelet quantile normalization method (subsection II-C)^1^. In section III, we describe the validation datasets and methodology: we briefly present the datasets used for benchmarking (III-A), the performance metrics (III-B), and the parameters used in the comparison (III-C). In section IV we discuss the results and present potential application to anesthesia monitoring. Finally, we present our conclusions in section V.

## II. Methods

### A. Discrete Wavelet Transform

We decompose the EEG signal *x* into time-frequency components using the discrete wavelet transform (DWT). The DWT coefficients are obtained by decomposing the signal onto an orthogonal basis obtained by dilating (scaling) and translating in time a wavelet function *ψ*(*t*) and scaling function *ϕ*(*t*) [32], [33]. The DWT can be computed efficiently with Mallat’s pyramidal algorithm, implemented by means of a hierarchical set of subband filters [34].

The DWT decomposition of the signal *x*(*t*) to the level *M* is defined as [32]:

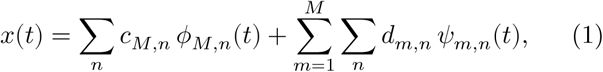

where

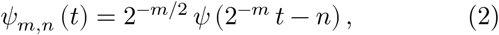

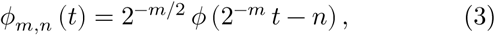

and *c*_*m,n*_ = ⟨*x, ϕ*_*m,n*_⟩ and *d*_*m,n*_ = ⟨*x, ψ* _*m,n*_⟩ are the approximation and detail coefficients at level *m*. The scalar product is defined by ⟨*f, g*⟩ = ∑_*t*∈𝕫_ *f*(*t*) *g*(*t*).

The DWT conserves energy so that:

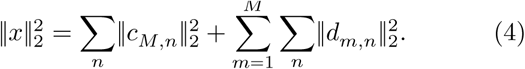

### B. Denoising by Wavelet Thresholding

Wavelet denoising methods rely on the assumption that artifactual components can be well isolated when the signal is represented in a wavelet basis. Denoising is performed by identifying and removing the wavelet coefficients associated with artifacts. For that goal, the signal is first decomposed onto a wavelet basis via DWT. Then, the wavelet coefficients are either removed or attenuated by a thresholding procedure. Finally, the corrected signal is recovered by inverting the wavelet transform.

The main challenge of this approach is therefore the selection of the appropriate threshold. While this could be done by manual inspection of the wavelet coefficients, this is unpractical in most applications. Various approaches for the automatic selection of thresholds of wavelet denoising have been proposed [35]–[38]. A common choice is the universal threshold [39] defined as:

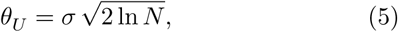

where σ is the standard deviation of the wavelet coefficients *w* and *N* is the number of coefficients. The value of σ can be estimated from the data using the median absolute deviation (MAD) estimator σ = *k* ⋅ median (|*w*|) with *k* ≈ 1.4826 [35].

Threshold values can be calculated separately for each DWT level. This procedure extends the universal threshold (eq. 5) to signals which are only weakly stationary and are correlated in time [38]. This is relevant in EEG artifact removal applications where stationarity is not guaranteed [40], [41]. In that case, eq. 5 becomes 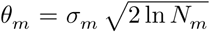 where σ_*m*_ and *N*_*m*_ refer to the wavelet coefficients at level *m*.

The thresholding procedure is usually carried out by applying a hard or a soft thresholding function on the wavelet coefficients *w*, defined as follows:

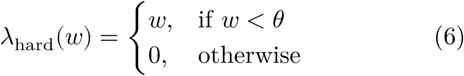

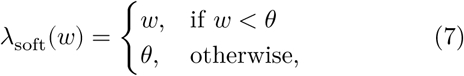

where *θ* is the chosen threshold (see Fig. 1).

**Fig. 1.**
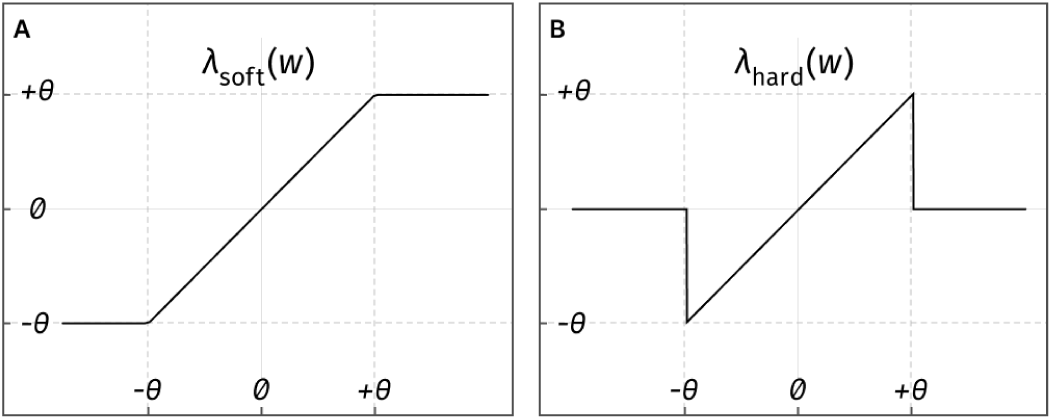
Threshold functions. **A** Soft thresholding (eq. 7). **B** Hard thresholding (eq. 6).

An alternative approach is the surrogate-based artifact removal method (SuBAR) [40], where time-varying thresholds are defined by considering the statistical deviation computed over an ensemble of surrogate time series. The surrogates are randomly generated via the iterative amplitude-adjusted Fourier transform (IAAFT) algorithm [42], [43] to produce stationary time series which have the same distribution as the original signal and a similar power spectrum. This method effectively amounts to detecting deviations from stationarity in the EEG signal and correct them via thresholding of the wavelet coefficients with the function λ_soft_ (eq. 7).

### C. Wavelet Quantile Normalization

As an alternative to wavelet thresholding, we develop here a novel renormalization technique for the wavelet coefficients which allows to adaptively attenuate artifactual components. The main idea of the method is that in most monitoring contexts the brain rhythm does not change dramatically during short time intervals (e.g. < 1 s). For example, in clinical monitoring of anesthesia brain activity is mostly controlled by hypnotic drugs. These drugs take effect gradually, producing brain waves patterns with a timescale of minutes [1]. We can thus consider that the distribution of energy across frequency bands is continuous and that there are no abrupt changes in the power of a band. We do not expect such continuity for transient brain activity, such as the response to an external stimulus [6]. Applying the proposed method in those situations would be inappropriate as it would probably attenuate genuine brain activity. In the following, we shall focus on continuous monitoring of the brain, where changes in the activity happen gradually. Under this continuity condition, the statistical characteristics such as the distribution of energy across frequencies of a short EEG fragment will be similar to the signal immediately preceding or following it.

The WQN method works as follows. First, we localize artifacts in the EEG signal (Fig. 2A) with traditional methods, such as thresholding of the signal power to detect non-physiological levels or using signal from auxiliary sensors (accelerometers, EMG, EOG) [44]. For each artifact, we construct a reference signal by considering some short portions of the EEG immediately before and/or after the occurrence of the artifact (Fig. 2B). Both the artifact and the reference segments are then decomposed via an *M*-level DWT (Fig. 2C), obtaining the coefficients 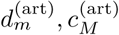 and 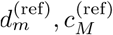. For uniformity of notation, we denote all coefficients as *w*_*m*_ = *d*_*m*_ for *m* = 1, …, *M* and *w*_*M*+1_ = *c*_*M*_.

**Fig. 2.**
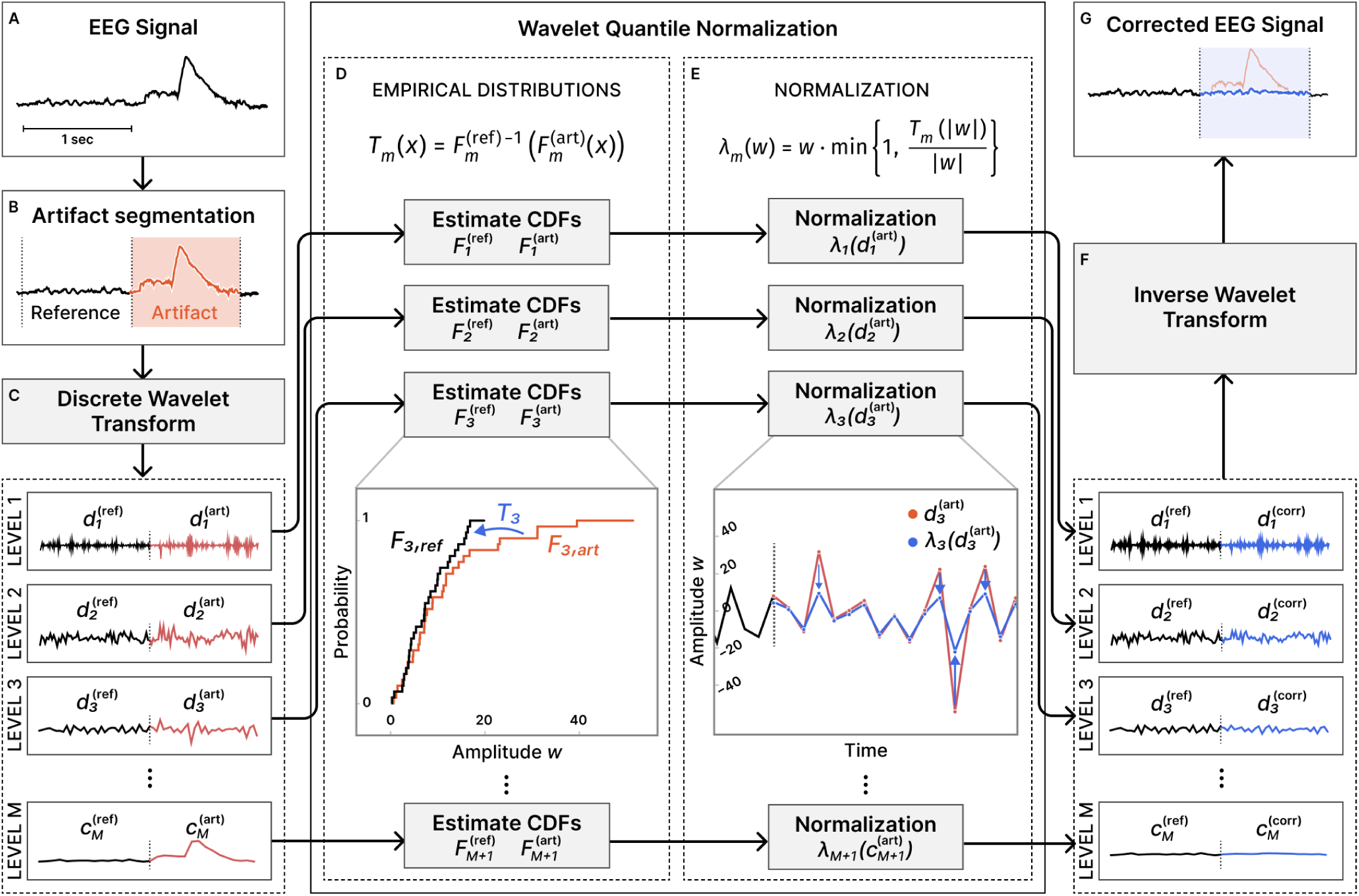
Diagram of proposed artifact removal method. **A** Input EEG signal (single-channel). **B** Artifact segmentation. **C** Decomposition with DWT on *M* levels. **D** Estimation of empirical cumulative density functions and transformation *T*_*m*_ (eq. 9). **E** Normalization of wavelet coefficients (eq. 10). **F** Reconstruction from corrected wavelet coefficients (eqs. 11–13). **G** Artifact-corrected EEG signal.

For each level *m* = 1, …, *M* + 1, we compute the empirical cumulative density function (CDF) 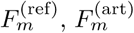 of the amplitude of the wavelet coefficients 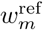 for reference and artifact (Fig. 2D) defined by

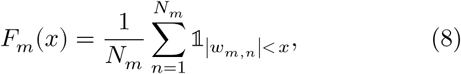

where 𝟙 is the indicator function. We can transform the wavelet coefficients 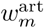 of the artifacted segment so that their distribution of amplitude will match that of the reference segment. This mapping is provided by the transformation *T*_*m*_ defined as

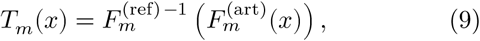

where 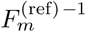 indicates the inverse of 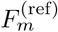. *T*_*m*_ transforms the wavelet coefficients 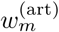 so that their amplitude distribution (and thus equivalently their energy distribution) becomes identical to that of 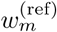 (see appendix I). This transformation adaptively attenuates high-power artifactual coefficients. Since we are only interested in reducing the power of artifacts, we restrict the normalization of coefficients to prevent amplification. Thus, we define the normalization function

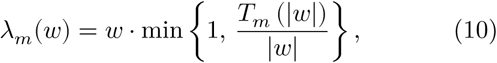

which maps a coefficient *w* from 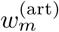 to its possibly attenuated value (Fig. 2E). Due to the energy conservation of the wavelet transform (eq. 4), attenuation of the wavelet coefficients leads to a reduction in the energy of specific time-frequency components of the original signal.

We define the corrected coefficients as:

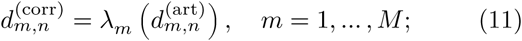

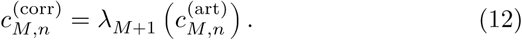

Finally, we reconstruct the corrected version of the artifacted segment by inverting the DWT as in eq. 1, using the corrected coefficients (Fig. 2F):

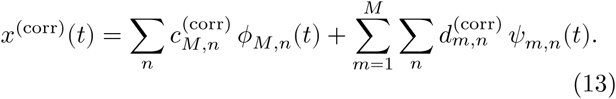

This algorithm can be applied to every artifacted signal interval to recover a corrected EEG (Fig. 2G).

## III. Validation

### A. Datasets for benchmarking

To validate the proposed method, we compare its performances on both unmodified and semi-simulated EEG recordings using publicly available datasets [44]–[46]. Each dataset comprises an artifacted EEG signal and a corresponding reference EEG signal used as ground truth. Artifact detection labels are given as input to the different algorithms.

#### 1) Physiobank Motion Artifacts dataset [44], [47]

This dataset contains 24 EEG recordings. The EEG signal was acquired by two closely placed electrodes producing highly correlated signals. One of the electrodes was then moved mechanically at regular time intervals, producing artifacts, while the other was kept still. Artifact removal can then be applied on the signal produced by the contaminated electrode, using the other channel as a ground truth reference. We filtered both channels in the range 0.1 Hz to 100 Hz with a Butterworth forward-backward filter of order 4 and down sampled to 256 Hz. Artifact detection was performed by thresholding the difference between the reference and the artifacted electrode. Recordings 10, 20, 21 were partially excluded after visual inspection revealed artifacts in the reference channel.

#### 2) Semi-simulated EEG/EOG [45]

This dataset contains 55 recordings from 19 EEG electrodes (200 Hz) and an equal number of EOG signals. The EOG signals are combined linearly to the EEG signal [45] to produce artifacted recordings. We modified the dataset to obtain 30 seconds recordings where the central 10 seconds are corrupted by EOG and marked as an artifact. Recording 36 was excluded after visual inspection.

#### 3) Denoise-Net [46]

This dataset includes thousands of 2-second epochs of EEG, EOG and EMG recorded at 256 Hz. We combined the clean EEG epochs with EOG and EMG to produce simulated datasets with 3400 partially artifacted epochs. These epochs were produced by summing one second artifact signal (EOG, EMG, or EOG+EMG) to the second half of the clean epoch (which was consequently marked as artifact). The amplitude of the artifact signal was rescaled to obtain different SNR values, in the range −20 dB to 5 dB (Fig. 3). In this dataset, the EEG signal of the artifacted epochs is dominated by either EOG or EMG but also contains the background brain activity. Thus the simulated partially-artifacted epochs are imperfect, as they may combine not only the artifact signal but also the brain activity from different subjects.

**Fig. 3.**
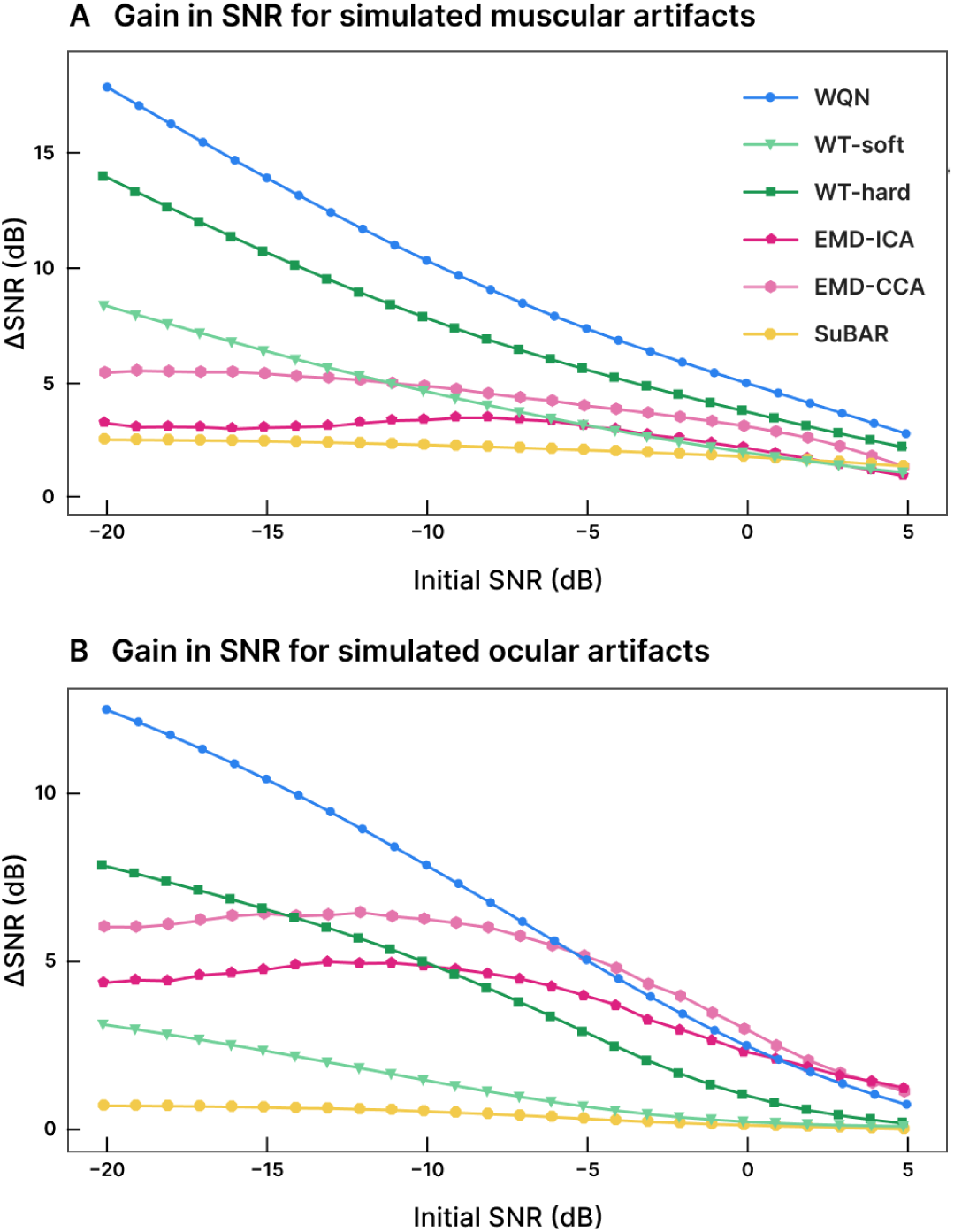
Noise reduction (ΔSNR) for varying level of simulated signal-to-noise ratio (SNR). **A** Simulated EMG dataset. **B** Simulated EOG dataset.

### B. Performance metrics

We employed different metrics to evaluate the performances of the artifact removal methods. Benchmarks were run with Python 3.9.7 on a Linux workstation equipped with an Intel Xeon Gold 5218R CPU (2.10 GHz).

#### 1) Change in signal to noise ratio

We define the metric ΔSNR as the change in the signal to noise ratio (SNR) before and after artifact removal. We only consider the signal epochs which were labeled as artifacted. The calculation of ΔSNR is defined as [21]:

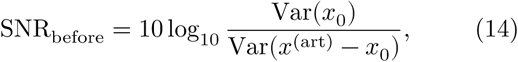

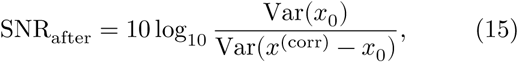

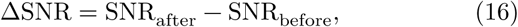

where Var(⋅) is the variance estimator, *x*_0_ is the ground truth signal, *x*^(art)^ is the artifacted signal, and *x*^(corr)^ is the signal obtained after artifact removal.

#### 2) Normalized mean squared error

The normalized mean squared error (NMSE), in decibels, is defined as:

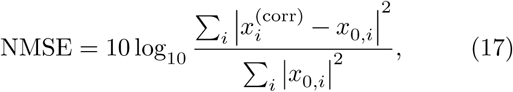

where *x*_*i*_ is the i-th sample of the signal *x*. Similarly to the definition of ΔSNR, we only consider signal epochs which are labeled as artifacted.

#### 3) Change in correlation

We defined the change in correlation Δ*R* before and after the artifact removal as:

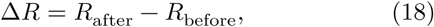

where *R*_before_ and *R*_after_ are the Pearson correlation coefficients between the ground truth and the signal before and after artifact removal respectively.

#### 4) Execution time

The execution time (see Table II) was calculated using a sample of duration 30 s at 256 Hz taken from the Physiobank dataset [44]. Each method was repeatedly applied to the same EEG sample, selecting the fastest run out of 10. All methods except SuBAR were implemented in Python and included in the published code. The original Matlab implementation provided by the authors was used for the SuBAR method [40].

### C. Methods parameters

We compare (Table I) our proposed wavelet quantile normalization method against the classical wavelet thresholding and the SuBAR method [40]. We implement the wavelet thresholding as described in section II-B, using the universal threshold defined in equation 5 separately for each decomposition level. Estimation of the standard deviation σ_*m*_ values is obtained by the MAD estimator σ_*m*_ = 1.4826⋅median (|*w*_*m*_|). We report the results for both hard and soft thresholding functions. The SuBAR method is applied on 30 s windows with no overlapping, using 1000 surrogates. For all tested methods, we used the symlet wavelet family with 5 vanishing moments [32]. In all cases, the correction was only applied on regions which were labeled as artifactual. EMD-CCA and EMD-ICA were implemented according to [21]. Up to 10 intrinsic mode functions were obtained from EMD and used as input for the CCA or ICA source separation. Artifactual sources were removed according to the methodology described in [21]: sources which, upon removal, increased the correlation to the reference signal were considered artifactual and thus suppressed. While this method cannot be employed in practice, since the reference signal is unknown, it ensures that the effectiveness of artifact removal is independent of the source classification strategy.

**TABLE I.** Result on validation on different datasets for the proposed WQN method, wavelet hard thresholding (WT-hard), wavelet soft thresholding (WT-soft), SuBAR, EMD-ICA, and EMD-CCA methods. ΔSNR and NMSE values are specified in dB. The best performance for each metric is highlighted in black.

| Dataset | Method | ΔSNR | NMSE | ΔR |
| --- | --- | --- | --- | --- |
| Motion Artifacts [44] | WQN (ours) | **16.57** | **0.66** | **0.51** |
|  | WT-hard (eq. 6) | 13.25 | 4.05 | 0.43 |
|  | WT-soft (eq. 7) | 7.60 | 9.69 | 0.22 |
|  | SuBAR [40] | 0.94 | 16.29 | 0.02 |
|  | EMD-ICA [21] | 5.91 | 11.33 | 0.19 |
|  | EMD-CCA [21] | 5.56 | 11.69 | 0.18 |
| Semi-simulated EOG [45] | WQN | **6.13** | **-5.80** | **0.06** |
|  | WT-hard | 3.32 | -2.97 | 0.03 |
|  | WT-soft | 1.55 | -1.22 | 0.01 |
|  | SuBAR | 1.72 | -1.39 | -0.01 |
|  | EMD-ICA | 2.08 | -1.76 | 0.03 |
|  | EMD-CCA | 2.35 | -2.03 | 0.03 |
| Denoise-Net [46] EMG | WQN | **17.84** | **2.67** | **0.46** |
|  | WT-hard | 13.90 | 6.48 | 0.34 |
|  | WT-soft | 8.28 | 11.85 | 0.18 |
|  | SuBAR | 2.44 | 17.61 | 0.04 |
|  | EMD-ICA | 3.16 | 16.91 | 0.10 |
|  | EMD-CCA | 5.36 | 14.95 | 0.14 |
| Denoise-Net [46] EOG | WQN | **12.48** | **10.41** | **0.24** |
|  | WT-hard | 7.81 | 13.95 | 0.17 |
|  | WT-soft | 3.08 | 18.39 | 0.05 |
|  | SuBAR | 0.66 | 20.82 | 0.00 |
|  | EMD-ICA | 4.36 | 17.34 | 0.15 |
|  | EMD-CCA | 6.00 | 15.94 | 0.19 |
| Denoise-Net [46] EOG+EMG | WQN | **13.47** | **9.20** | **0.25** |
|  | WT-hard | 8.46 | 13.13 | 0.16 |
|  | WT-soft | 4.05 | 17.28 | 0.06 |
|  | SuBAR | 1.24 | 19.90 | 0.01 |
|  | EMD-ICA | 2.22 | 18.59 | 0.08 |
|  | EMD-CCA | 2.95 | 17.99 | 0.10 |

## IV. Results and Discussion

### A. Artifact removal effectiveness

To validate the effectiveness of the WQN method we benchmarked it against alternative artifact removal algorithms which are suitable for single-channel EEG recordings: wavelet thresholding (with both soft and hard thresholding functions), EMD-ICA, EMD-CCA [21], and surrogate-based artifact removal (SuBAR) [40]. We included artifacts of different nature (movement, ocular, muscular) using datasets of semi-simulated and real EEGs (section III-A), where we used a ground truth signal to evaluate the performances of artifact removal algorithms. We quantified the reduction of artifactual components using the normalized mean squared error (NMSE), the gain in SNR (ΔSNR), and the improvement in correlation (ΔR), obtained by comparing the output of the artifact removal algorithms to the ground truth signal (section III-B). We note that in some cases, an algorithm can show an improvement in terms of ΔSNR and NMSE but at the same time destroy the structure of the signal, losing the information content of the data. This is particularly the case for high-power artifacts. To avoid this effect, we consider the improvement in correlation (ΔR) as well as NMSE and ΔSNR. Since a degradation of the signal will coincide with a lower correlation to the ground truth, we consider that a simultaneous improvement in ΔSNR and ΔR is a solid indication of reliable artifact removal.

We found that WQN performed better among the tested methods in terms of SNR and NMSE, producing the highest increase to signal to noise ratio (ΔSNR) and the lowest normalized mean squared error on all datasets. WQN also outperformed the other methods in terms of improvement of the correlation, confirming absence of information loss. For the tested data, the performances of the SuBAR method were lower to those of wavelet thresholding. This result can be understood because the SuBAR was designed to remove short, isolated artifacts and was reported to be ill-suited for large EOG artifacts [40]. EMD-ICA and EMD-CCA showed better performances in removing ocular and motion artifacts, but were significantly outperformed by wavelet-based methods in the case of high-frequency muscular artifacts. This result confirms that ICA and similar BSS methods might not be optimal in removing EMG activity [13], [48]. Our tests also revealed problems in the convergence of the FastICA algorithm when applied to the 2-second EEG samples from the Denoise-Net datasets, highlighting the limitations of this method for short time windows. The results of the benchmark are summarized in Table I.

We then investigated how the performances depend on the initial signal-to-noise level. To test this, we rescaled the amplitude of the artifacts in the Denoise-Net datasets (sec. III-A.3) to obtain SNRs between −20 dB (large, powerful artifacts) and 5 dB (small artifacts). In Figure 3 we show the ΔSNR for the various initial SNR values on muscular and ocular artifacts. WQN consistently outperformed the other methods in the case of muscular artifacts, showing higher ΔSNR for all initial SNR levels. In this case, SuBAR outperformed soft wavelet thresholding for weak muscular artifacts (Fig. 3A). For ocular artifacts, EMD-based methods and WQN showed the best performances, with WQN significantly outperforming other methods for low SNRs (< −10 dB).

In Figure 4 we show three examples of corrected artifacts obtained with the WQN method: motion artifact (Fig. 4A), eye blink (Fig. 4B), and muscular activity (Fig. 4C). The results show a significant reduction of the artifacts without distortion of the EEG signal, highlighting the flexibility and robustness of the proposed method.

**Fig. 4.**
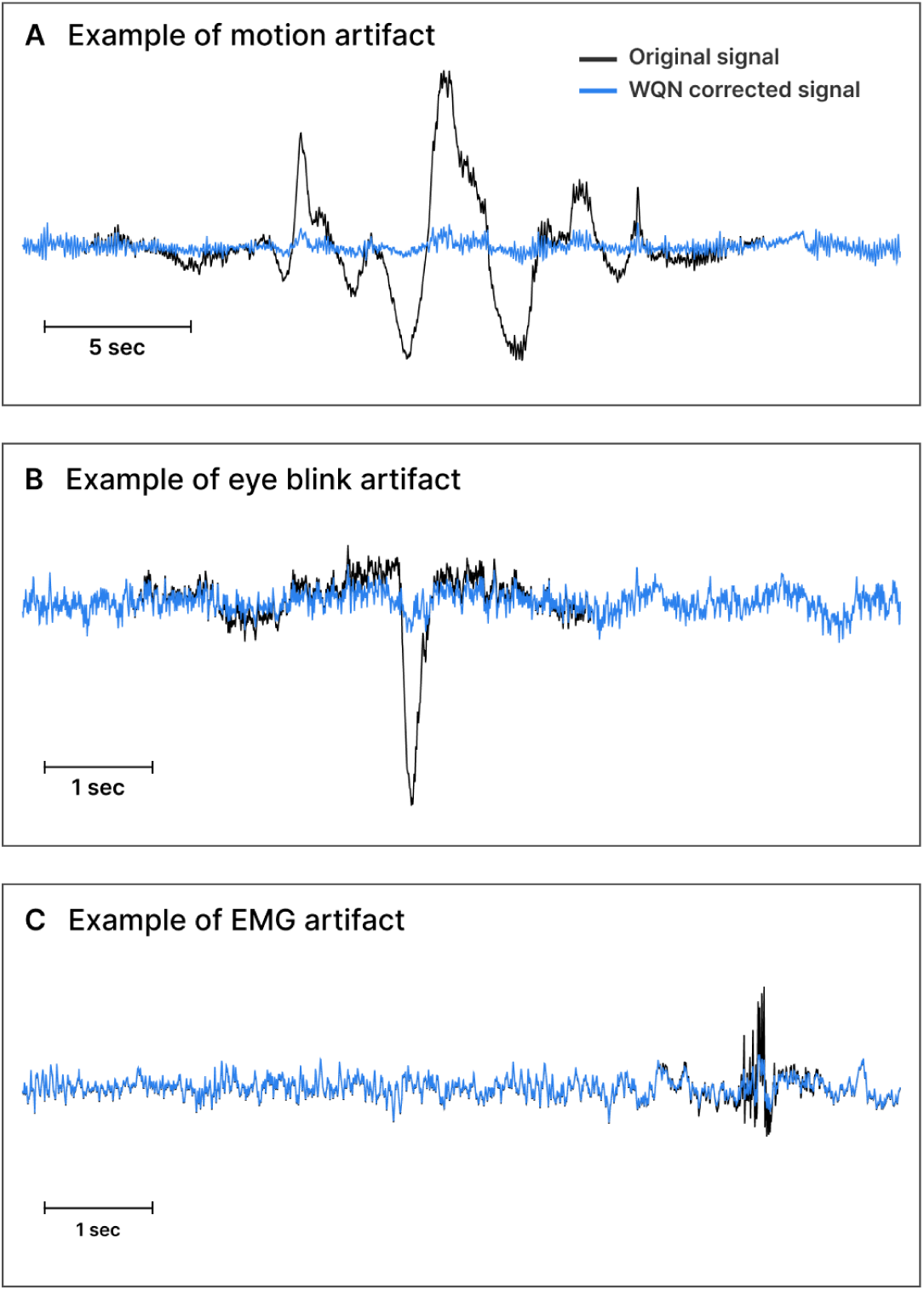
Example of artifact removal via WQN (original signal in black, corrected signal in blue). **A** Motion artifact. **B** Eye blink artifact. **C** Muscular artifact (EMG). (The samples were obtained from datasets [44], [55], [56].)

### B. Computational complexity

We consider a single-channel EEG signal of length *N* samples. The computational complexity of a M-level discrete wavelet transform is 𝒪(*MN*) [34], [51]. In wavelet thresholding algorithms, the threshold value is typically fixed beforehand or can be determined in 𝒪(*N*) time based on the signal statistics (such as the universal thresholding presented in equation 5). Thus, the leading term in computational complexity is 𝒪(*MN*) due to the DWT decomposition.

A similar result holds for the proposed WQN algorithm. The the M-level DWT can be performed in 𝒪(*MN*) operations. For an artifact of length *N*_art_, the empirical cumulative distribution is obtained by sorting the coefficients with a cost of 𝒪(*N*_art_ log *N*_art_). The total computational complexity can thus be given as 𝒪(*MN* +*K*_art_*N*_art_ log *N*_art_) for *K*_art_ artifacts of maximum length *N*_art_. Since the number and length of artifacts is typically limited, the computational complexity will be dominated by the 𝒪(*MN*) term. In practice, the WQN algorithm can be faster than WT since the DWT has to be computed only on short fragments of the signal around the artifact.

In the SuBAR method [40] the leading term of computational complexity comes from the generation of Fourier surrogates via Iterative Amplitude Adjusted Fourier Transform (IAAFT) [42]. Each iteration of the IAAFT algorithm requires the computation of Fourier transform and sorting of the coefficients, with both operations requiring 𝒪(*N* log *N*) time. The generation of *K*_su_ surrogates has thus a computational complexity of 𝒪(*K*_su_ *K*_it_ *N* log *N*) where *K*_it_ is the number of iterations of the IAAFT algorithm.

EMD-based methods such as EMD-ICA and EMD-CCA require the decomposition of the signal as a sum of intrinsic mode functions (IMFs). Although EMD is commonly reported to be computationally demanding [52], its order of complexity is 𝒪(*N* log *N*) [52], [53]. Moreover, if only the first *M* IMFs are considered, the complexity becomes 𝒪(*MN*) [51], [52], making it equivalent to the M-level DWT. In our empirical measures of the CPU time (Table II), the EMD-based algorithms remain significantly slower than wavelet decomposition (∼300 ms versus 3 ms respectively). ICA is commonly implemented via the FastICA algorithm which has computational cost 𝒪(*K*_it_*M* ^2^*N*) [50], [51] where *K*_it_ is the number of algorithm iterations. CCA can be efficiently implemented by singular value decomposition with complexity 𝒪(*M* ^2^*N*) and typically outperforms ICA [21], [54].

**TABLE II.**
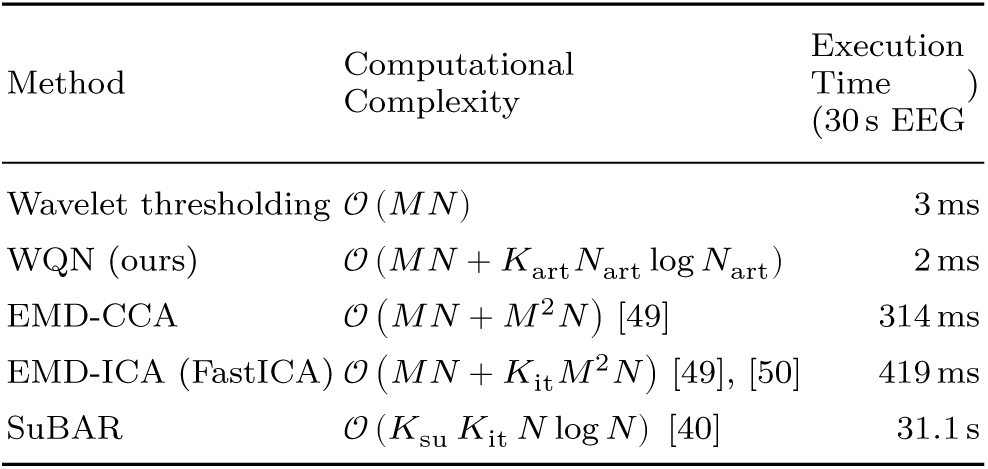
Computational cost of different algorithms. The theoretical computational complexity is reported for a signal of length *N* using *M* levels of decompositions/instrinsic mode functions. The execution time is the empirical CPU time required to analyze 30 s EEG signal at 256 Hz.

| Method | Computational Complexity | Execution Time (30 s EEG) |
| --- | --- | --- |
| Wavelet thresholding | $\mathcal{O}(MN)$ | 3 ms |
| WQN (ours) | $\mathcal{O}(MN + K_{\text{art}} N_{\text{art}} \log N_{\text{art}})$ | 2 ms |
| EMD-CCA | $\mathcal{O}(MN + M^2 N)$ [49] | 314 ms |
| EMD-ICA (FastICA) | $\mathcal{O}(MN + K_{\text{it}} M^2 N)$ [49], [50] | 419 ms |
| SuBAR | $\mathcal{O}(K_{\text{su}} K_{\text{it}} N \log N)$ [40] | 31.1 s |

We then compared the performances of practical implementations of the algorithms by measuring the CPU time required to analyze a 30-second EEG sample taken from the Physiobank dataset [44]. WQN and wavelet thresholdings showed similar performances (2 ms and 3 ms respectively) and were superior to EMD-based and SuBAR algorithms (314 ms for EMD-CCA, 419 ms for EMD-ICA, and 31 s for SuBAR).

In conclusion, we found that the computational cost of the WQN method is comparable to that of wavelet thresholding and that both are significantly faster than EMD and surrogate-based method. The computational complexity and the CPU execution time for each algorithm are summarized in Table II.

### C. Application to anesthesia monitoring

We present an application of WQN to the monitoring of general anesthesia. In this context, spectral measures of the EEG such as the spectral edge frequency (SEF) or the alpha-to-delta ratio (ADR) are routinely used by anesthesiologists to control the depth of anesthesia. Artifacts caused by inadvertent motion of the electrodes can alter the signal spectrum and make these measures unreliable (Fig. 5A-B). Depending on the duration of the time window on which the spectrum is calculated, a single artifact can affect the output of the monitoring for up to one minute. In figure 5 we present an example of the EEG spectrogram of a patient during the maintenance phase of general anesthesia, showing how artifacts (purple arrows) can negatively affect the calculation of the SEF95 (Fig. 5B). WQN can be used to recover a corrected spectrum (Fig. 5C) and to obtain a more robust estimation of spectral measures such as the SEF, improving the reliability of anesthesia monitoring.

**Fig. 5.**
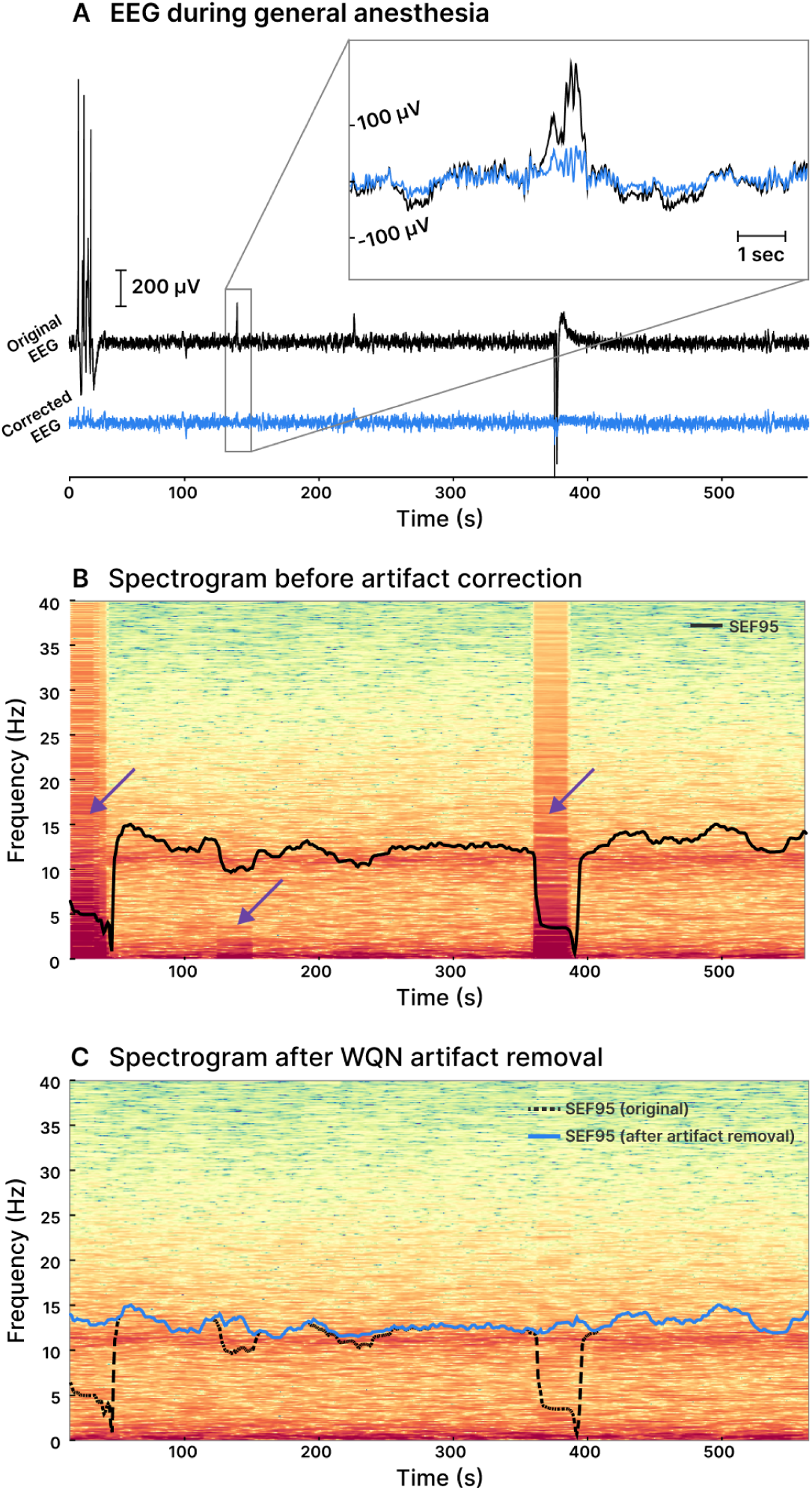
Example of artifact removal for EEG during anesthesia. **A** Original EEG (black line) and WQN-corrected EEG (blue line) of a patient under general anesthesia. **B** Spectrogram and Spectral Edge Frequency at 95% (SEF95, black line). Artifacts are visibly contaminating the spectrum (purple arrows) and corrupting the SEF95 calculation. **C** Spectrogram and SEF95 after artifacts were removed using Wavelet Quantile Normalization. The EEG signal was extracted from the VitalDB dataset [57].

### D. Limitations of the proposed method

The WQN method assumes gradual change of the brain activity, suppressing patterns which are associated to abrupt power changes. While this proved to be effective for several types of artifacts (Table I), the assumption cannot be extended to contexts where artifacts are associated to other transient activity. An example are epileptic seizures which provoke sudden changes in the EEG power due to abnormal brain activity and are simultaneously associated to muscular or movement artifacts. Application of WQN in these cases would attenuate the epileptic patterns together with the artifactual components, potentially causing loss of important clinical information. Overcoming this limitation would require a spatio-temporal model to separate abnormal patterns of epileptic and artifactual origin [58], an issue closely related to the detection and classification of epileptic seizures.

## V. Conclusions

In this work we presented and validated an artifact removal algorithm which can operate on single-channel EEG. The proposed WQN method is data-driven and requires no auxiliary input, parameter tuning or any human intervention. In our tests, WQN consistently outperformed comparable algorithms when applied on artifacts of different nature (Table I, Fig. 3 and 4) and for different signal-to-noise ratios. Moreover, we showed that the proposed algorithm has a similar computational cost to wavelet thresholding algorithms and that its performances are compatible with real-time applications. The characteristics of the method make it particularly suited for anesthesia monitoring and other special care unit applications where only few electrodes are available and analysis has to be performed in real-time.

We presented an example of application to general anesthesia (Fig. 5), showing how the WQN method could be used to improve the reliability of EEG monitoring. Future work includes the development and integration of an artifact detection algorithm, making WQN a fully automated technique to reduce artifacts in unsupervised EEG devices.

## Data Availability

All data produced in the present study are available upon reasonable request to the authors

## Appendix I Probability transformation

Consider a random variable *X*, with probability density function *f*_*X*_

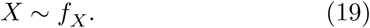

We would like to generate random variables distributed as *f*_*Y*_, i.e. we want to find a monotonic transformation *T* such that

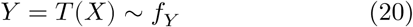

We obtain *T* in two steps. First, using the probability integral transform, we map *X* to a uniform random variable *U*

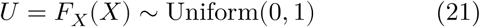

where *F*_*X*_ is the cumulative density function of *X*. Then, with the inverse integral transform we map *U* to *Y*

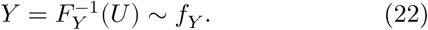

Combining the two steps, we finally define

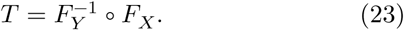

The validity of this result can be easily verified by checking that the cumulative density function of *T* (*X*) is indeed *F*_*Y*_ :

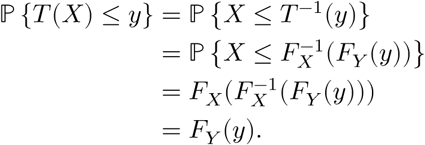

## Acknowledgments

The authors would like to thank Mario Chavez for kindly providing the Matlab implementation of the SuBAR algorithm. We also thank Pierre Lozeron and Nathalie Kubis for discussion.

This paper includes material from the authors’ pending patent “Automated EEG signal analysis and construction of a probability map to predict the sensitivity to general anesthesia during induction to prevent over-sedation”.

## Footnotes

1 The Python code implementing the WQN method and benchmarking procedure will be available at https://github.com/holcman-lab/wavelet-quantile-normalization (DOI 10.5281/zenodo.4783450)

